# Association of Pulmonary Artery Compliance and Adverse Cardiac Events

**DOI:** 10.1101/2025.06.19.25329923

**Authors:** Louisa A Mounsey, Sophie M Nemeth, Leah B Kosyakovsky, Jenna N McNeill, Michael H Picard, Eugene V Pomerantsev, Juhi K Parekh, Noah Schoenberg, David Furfaro, Cyrus Kholdani, Jennifer E Ho

## Abstract

**Introduction:** Pulmonary artery compliance (PAC), the ratio of stroke volume to pulmonary artery pulse pressure, reflects the pulsatile component of right ventricular afterload. While lower PAC is associated with right heart failure (HF) and death in patients with pulmonary arterial hypertension, the association of PAC with clinical outcomes among individuals with a broader range of cardiopulmonary comorbidities is unclear.

**Methods:** We examined consecutive ambulatory and hospitalized patients undergoing clinically indicated right heart catheterization at a single center between 2005 and 2016. Multivariable Cox models were used to investigate the association of PAC with clinical outcomes including HF hospitalization and mortality. Analyses were stratified by presence and absence of pulmonary hypertension (PH) and by PH hemodynamic subtype.

**Results:** Among 7966 patients (mean age 63 years, 39% women), median PAC was 3.29 (IQR 2.19, 4.70) mL/mmHg. PAC was significantly inversely associated with mortality (HR 0.59 per 1-SD higher PAC, 95% CI 0.56, 0.63) and HF hospitalization (HR 0.56, 95% CI 0.52, 0.59) across the whole sample and among those with PH (mortality: HR 0.69, 95% CI 0.64, 0.74, HF: HR 0.65, 95% CI 0.61, 0.70) and without PH (mortality: HR 0.74, 95% CI 0.66, 0.84, HF: HR 0.72, 95% CI 0.61, 0.85). Similarly, PAC was associated with mortality across all PH subtypes: precapillary PH (HR 0.56, 95% CI 0.48, 0.65), combined PH (HR 0.75, 95% CI 0.66, 0.86) and isolated post-capillary PH (HR 0.88, 95% CI 0.79, 0.98).

**Conclusions:** Among patients undergoing clinically indicated right heart catheterization, lower PAC is associated with adverse outcomes irrespective of presence or absence of PH. Our findings support potential clinical utility of PAC in risk stratification across a broad spectrum of cardiopulmonary disease.

## Introduction

Pulmonary hypertension (PH) results from pulmonary vascular remodeling leading to increased pulmonary vascular resistance (PVR), decreased pulmonary arterial compliance (PAC), and ultimately death from right heart failure. Hemodynamic characterization of PH aims to determine the driving pathophysiology of elevated pulmonary artery pressures and to identify high risk patients to allow for early implementation of therapeutic efforts.^1,2^ Risk stratification tools have predominantly utilized PVR to quantify right ventricular (RV) afterload, yet it is important to recognize that PVR estimates resistive, and not pulsatile afterload. By contrast, PAC, the ratio of stroke volume to pulmonary artery pulse pressure 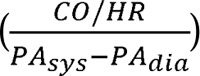 reflects the pulsatile component of RV afterload, and may serve as an early and complementary indicator of pulmonary vascular disease apart from PVR.^3,4^ Another derived hemodynamic variable, the resistance-compliance (RC) time constant (PAC X PVR), traditionally was felt to be constant across PH hemodynamic subtypes, yet more recently has been shown to differ between those with pre-vs post-capillary disease.^5–7^

Prior studies have examined both PAC and RC in specific populations such as those with pulmonary arterial hypertension, chronic thromboembolic pulmonary hypertension, and heart failure with preserved ejection fraction, yet few have evaluated their prognostic significance across a broader spectrum of cardiopulmonary disease and hemodynamic abnormalities.^8–10^ We leveraged a large cohort of patients who underwent clinically-indicated right heart catheterization (RHC) to 1) examine the association of PAC with adverse clinical outcomes including death and heart failure hospitalization, 2) assess the prognostic implication of PAC across PH hemodynamic subtypes, and 3) evaluate PAC and RC by PH hemodynamic subtype to place these parameters in context of other markers of disease severity.

## Methods

### Study sample

We examined consecutive ambulatory and hospitalized patients undergoing RHC at Massachusetts General Hospital between 2005 and 2016. For patients with multiple RHC procedures, only the initial test was included, resulting in a total of 10,306 patients. Exclusion criteria included acute myocardial infarction (MI) occurring on the same day as the procedure, cardiac arrest or shock within 24 hours of the procedure, presence of mechanical ventilation, presence of an intra-aortic balloon pump, history of heart or lung transplant, complicated adult congenital heart disease, history of valve replacement, and dialysis (n=887 excluded) (**Figure 1**). *Additional exclusion criteria included missing key clinical covariates (n= 484),* patient identifier variables (n= 398), and hemodynamic parameters (n= 571), leaving a final study sample of 7966 for analysis. The study was approved by the local institutional review board.

**Figure 1:**
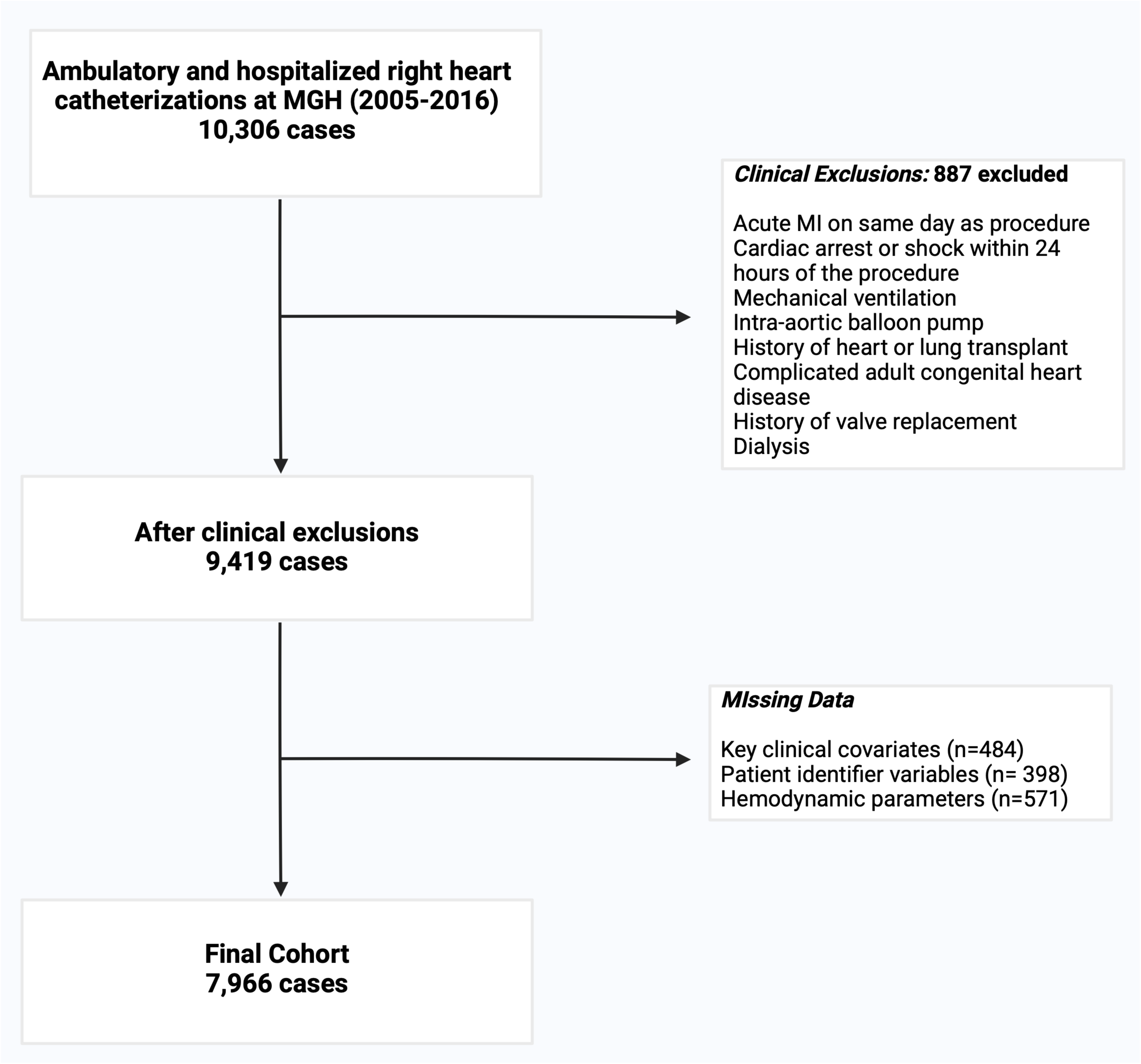
Flowchart demonstrating application of exclusion criteria.

### Clinical and hemodynamic assessment

Demographics, including age, sex, body mass index (BMI), and comorbidities, including history of MI, heart failure (HF), hypertension, diabetes, chronic kidney disease, lung disease, and obstructive sleep apnea/ obesity hypoventilation syndrome (OSA/OHS), were abstracted from the medical record at the time of RHC. As previously described, echocardiographic data within one year of RHC were reviewed with abstraction of quantitative and qualitative parameters.^11^ Echocardiographic data were available for 6478 patients (81%).

Hemodynamic variables recorded at RHC include resting systemic blood pressure, heart rate, right atrial (RA) pressure, pulmonary artery (PA) systolic, diastolic, and mean pressure, and mean pulmonary capillary wedge pressure (PCWP). Cardiac output was measured via the thermodilution method. Indirect Fick cardiac output was substituted for 548 patients with unavailable thermodilution values. Non-physiologic values (i.e. zero or negative) were set to missing. Pulmonary vascular resistance was calculated by 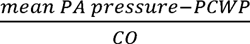, PAC by 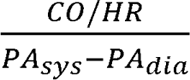, and resistance-compliance (RC) time constant by PVR X PAC.

Patients with a mean PA pressure >20 mmHg were considered to have PH. PH subgroups were defined as pre-capillary (PVR ≥ 2 Wood Units (WU) and PCWP ≤15 mmHg), isolated post-capillary pulmonary hypertension (IpcPH) (PVR < 2 WU and PCWP >15mmHg), or combined pre-capillary and post-capillary pulmonary hypertension (CpcPH) (PVR >2 and PCWP >15 mmHg).^2^ There were 585 patients with a mean PA >20 mmHg who did not meet criteria for PH subgroup assignment (i.e. PVR <2 WU and PCWP <15 mmHg). They were included in analyses by presence versus absence of PH, but not in analyses by PH hemodynamic subtype.

### Clinical outcomes

The primary outcomes were all-cause mortality and HF hospitalization. All-cause mortality was ascertained using the National Social Security Death Master Index and hospital records, abstracted on June 10, 2020. The precise dates of deaths that occurred between June 10, 2017 and the date of abstraction (a 3-year period) are protected nationally due to confidentiality purposes. Thus, these death events were imputed as occurring midway through this period, December 10, 2018, for standardization (n=242 individuals). HF hospitalization was defined by an *International Classification of Diseases Ninth Revision* or *International Classification of Diseases Tenth Revision* (ICD-10) code for HF as primary discharge diagnosis or a current procedural terminology (CPT) code for heart transplantation or durable ventricular assist device. The follow up period was defined as time from right heart catheterization to chart abstraction or death.

### Statistical analysis

PAC was natural log-transformed to yield a normal distribution and scaled. Quartiles of log-transformed PAC were defined by a percentile-based approach to yield equal group sizes. The distribution of PVR was winsorized to limit effect of outliers on the analysis, setting the minimum PVR at 0.2 W.U and the maximum at 20 W.U. PVR and RC were natural log-transformed and scaled. Demographics, comorbidities, echocardiographic data, and hemodynamic data are displayed by PAC quartile with mean and standard deviation for numerical variables, unless otherwise noted, and count and percent for categorical variables.

Clinical correlates of PAC, including BMI, history of hypertension, diabetes mellitus, previous MI, chronic HF, chronic kidney disease, OSA/OHS, chronic lung disease, and N-Terminal Pro-B-Type Natriuretic Peptide (NT-proBNP), were analyzed using age- and sex-adjusted linear regression models. Stepwise regression analysis was utilized to determined multivariate correlates of PAC, with entry criteria set at p < 0.01 and retention at p < 0.05. RHC hemodynamic correlates of PAC were analyzed using Pearson partial correlation coefficients, adjusted for age and sex.

The Kaplan Meier method was used to estimate the probability of each primary outcome across PAC quartiles in the entire cohort, by presence and absence of PH, and by PH subgroup. Log rank tests with Benjamini-Hochberg adjustment for false discovery rate were used to compare outcomes by quartile. Multivariable Cox models were constructed to separately examine the association of PAC with all-cause mortality and HF hospitalization. Models were first adjusted for age and sex, with further addition of BMI, history of hypertension, diabetes mellitus, previous myocardial infarction, chronic HF, chronic kidney disease, OSA/OHS, and chronic lung disease. A third model additionally included PVR. The proportional hazards assumption was assessed using Schoenfeld residuals and time-dependent covariates were incorporated as appropriate.

The Wilcoxon rank sum test was used to compare PAC by presence or absence of PH. RC time constant was compared between all PH hemodynamic subgroup pairs with the Kruskal-Wallis rank sum test, and between each subgroup pair with *t* tests. To assess the prognostic utility of PAC in ascertaining the risk of all-cause mortality and HF hospitalization compared to other pulmonary vascular indices, we examined model discrimination using c-statistics for Cox models containing clinical variables as above, with and without the addition of PAC, PVR, and RC time constant.

Statistical analyses were performed using R version 4.1.2, save for the assessment of model discrimination, for which PROC PHREG in SAS Institute Inc (Cary, NC) was utilized.^12^

## Results

### Baseline characteristics

We studied 7,966 patients with mean age 63±13 years, 39% women, and mean BMI of 29±7 kg/m^2^. Cardiometabolic and pulmonary comorbidities were frequent, including 59% with hypertension, 32% with prior HF, and 16% with chronic lung disease. The median PAC was 3.29 mL/mmHg (IQR 2.19, 4.70). Baseline clinical and characteristics by PAC quartile are displayed in **Table 1**.

**Table 1.**
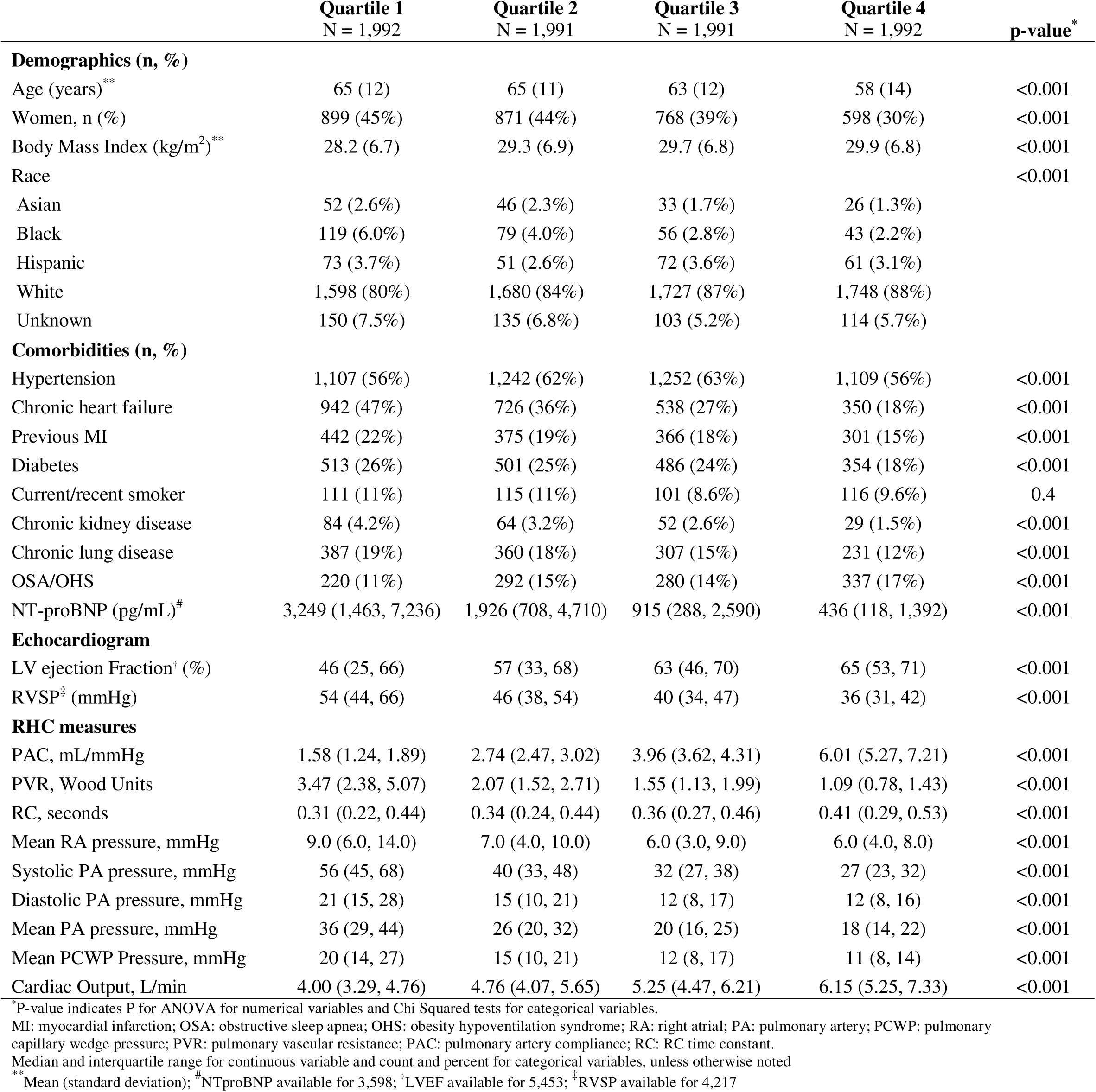
Clinical, echocardiographic, and hemodynamic measures across PAC quartiles.

|  | Quartile 1<br>N = 1,992 | Quartile 2<br>N = 1,991 | Quartile 3<br>N = 1,991 | Quartile 4<br>N = 1,992 | p-value* |
| --- | --- | --- | --- | --- | --- |
| **Demographics (n, %)** |  |  |  |  |  |
| Age (years)** | 65 (12) | 65 (11) | 63 (12) | 58 (14) | <0.001 |
| Women, n (%) | 899 (45%) | 871 (44%) | 768 (39%) | 598 (30%) | <0.001 |
| Body Mass Index (kg/m<sup>2</sup>)** | 28.2 (6.7) | 29.3 (6.9) | 29.7 (6.8) | 29.9 (6.8) | <0.001 |
| Race |  |  |  |  | <0.001 |
| Asian | 52 (2.6%) | 46 (2.3%) | 33 (1.7%) | 26 (1.3%) |  |
| Black | 119 (6.0%) | 79 (4.0%) | 56 (2.8%) | 43 (2.2%) |  |
| Hispanic | 73 (3.7%) | 51 (2.6%) | 72 (3.6%) | 61 (3.1%) |  |
| White | 1,598 (80%) | 1,680 (84%) | 1,727 (87%) | 1,748 (88%) |  |
| Unknown | 150 (7.5%) | 135 (6.8%) | 103 (5.2%) | 114 (5.7%) |  |
| **Comorbidities (n, %)** |  |  |  |  |  |
| Hypertension | 1,107 (56%) | 1,242 (62%) | 1,252 (63%) | 1,109 (56%) | <0.001 |
| Chronic heart failure | 942 (47%) | 726 (36%) | 538 (27%) | 350 (18%) | <0.001 |
| Previous MI | 442 (22%) | 375 (19%) | 366 (18%) | 301 (15%) | <0.001 |
| Diabetes | 513 (26%) | 501 (25%) | 486 (24%) | 354 (18%) | <0.001 |
| Current/recent smoker | 111 (11%) | 115 (11%) | 101 (8.6%) | 116 (9.6%) | 0.4 |
| Chronic kidney disease | 84 (4.2%) | 64 (3.2%) | 52 (2.6%) | 29 (1.5%) | <0.001 |
| Chronic lung disease | 387 (19%) | 360 (18%) | 307 (15%) | 231 (12%) | <0.001 |
| OSA/OHS | 220 (11%) | 292 (15%) | 280 (14%) | 337 (17%) | <0.001 |
| NT-proBNP (pg/mL)<sup>#</sup> | 3,249 (1,463, 7,236) | 1,926 (708, 4,710) | 915 (288, 2,590) | 436 (118, 1,392) | <0.001 |
| **Echocardiogram** |  |  |  |  |  |
| LV ejection Fraction<sup>†</sup> (%) | 46 (25, 66) | 57 (33, 68) | 63 (46, 70) | 65 (53, 71) | <0.001 |
| RVSP<sup>‡</sup> (mmHg) | 54 (44, 66) | 46 (38, 54) | 40 (34, 47) | 36 (31, 42) | <0.001 |
| **RHC measures** |  |  |  |  |  |
| PAC, mL/mmHg | 1.58 (1.24, 1.89) | 2.74 (2.47, 3.02) | 3.96 (3.62, 4.31) | 6.01 (5.27, 7.21) | <0.001 |
| PVR, Wood Units | 3.47 (2.38, 5.07) | 2.07 (1.52, 2.71) | 1.55 (1.13, 1.99) | 1.09 (0.78, 1.43) | <0.001 |
| RC, seconds | 0.31 (0.22, 0.44) | 0.34 (0.24, 0.44) | 0.36 (0.27, 0.46) | 0.41 (0.29, 0.53) | <0.001 |
| Mean RA pressure, mmHg | 9.0 (6.0, 14.0) | 7.0 (4.0, 10.0) | 6.0 (3.0, 9.0) | 6.0 (4.0, 8.0) | <0.001 |
| Systolic PA pressure, mmHg | 56 (45, 68) | 40 (33, 48) | 32 (27, 38) | 27 (23, 32) | <0.001 |
| Diastolic PA pressure, mmHg | 21 (15, 28) | 15 (10, 21) | 12 (8, 17) | 12 (8, 16) | <0.001 |
| Mean PA pressure, mmHg | 36 (29, 44) | 26 (20, 32) | 20 (16, 25) | 18 (14, 22) | <0.001 |
| Mean PCWP Pressure, mmHg | 20 (14, 27) | 15 (10, 21) | 12 (8, 17) | 11 (8, 14) | <0.001 |
| Cardiac Output, L/min | 4.00 (3.29, 4.76) | 4.76 (4.07, 5.65) | 5.25 (4.47, 6.21) | 6.15 (5.25, 7.33) | <0.001 |

\*P-value indicates P for ANOVA for numerical variables and Chi Squared tests for categorical variables.
MI: myocardial infarction; OSA: obstructive sleep apnea; OHS: obesity hypoventilation syndrome; RA: right atrial; PA: pulmonary artery; PCWP: pulmonary capillary wedge pressure; PVR: pulmonary vascular resistance; PAC: pulmonary artery compliance; RC: RC time constant.
Median and interquartile range for continuous variable and count and percent for categorical variables, unless otherwise noted
\*\* Mean (standard deviation); <sup>#</sup>NTproBNP available for 3,598; <sup>†</sup>LVEF available for 5,453; <sup>‡</sup>RVSP available for 4,217

### Clinical and hemodynamic correlates of PAC

In age- and sex-adjusted analyses, we found that lower PAC was associated with older age, female sex, and presence of comorbid conditions including history of HF, prior MI, diabetes, chronic kidney disease, and chronic lung disease (p ≤0.001 for all, **Table 2**). For example, individuals with HF had a lower PAC compared with those without HF (ß estimate −0.46, standard error 0.02, p <0.001). By contrast, higher BMI, hypertension and the presence of OSA/OHS were associated with higher PAC (p <0.001 for all). In a stepwise multivariable regression model, we found that age, sex, BMI, hypertension, history of HF, diabetes, chronic kidney disease, and chronic lung disease remained independently associated with PAC (**Table 2**).

**Table 2:** Models of clinical and right heart catheterization measure correlates of pulmonary artery compliance (PAC)

| Predictors | Age/sex adjusted model |  |  | Stepwise model |  |  |
| --- | --- | --- | --- | --- | --- | --- |
| | $\beta$ Estimate | Standard Error | p-value | $\beta$ Estimate | Standard Error | p-value |
| Clinical variables |  |  |  |  |  |  |
| Age | -0.22 | 0.01 | <0.001 | -0.22 | 0.01 | <0.001 |
| Sex (male) | 0.24 | 0.02 | <0.001 | 0.26 | 0.02 | <0.001 |
| BMI | 0.09 | 0.01 | <0.001 | 0.09 | 0.01 | <0.001 |
| Hypertension | 0.16 | 0.02 | <0.001 | 0.25 | 0.02 | <0.001 |
| Chronic heart failure | -0.46 | 0.02 | <0.001 | -0.48 | 0.01 | <0.001 |
| Previous MI | -0.13 | 0.03 | <0.001 | - | - | - |
| Diabetes | -0.08 | 0.03 | 0.001 | -0.09 | 0.03 | 0.001 |
| Chronic kidney disease | -0.32 | 0.06 | <0.001 | -0.20 | 0.06 | 0.001 |
| Chronic lung disease | -0.15 | 0.03 | <0.001 | -0.12 | 0.03 | <0.001 |
| OSA/OHS | 0.13 | 0.03 | <0.001 | 0.05 | 0.03 | 0.09 |
| NT-proBNP* | -0.42 | 0.01 | <0.001 | - | - | - |
|  | Correlation coefficient (r) |  | p-value |  |  |  |
| RHC measures |  |  |  |  |  |  |
| Mean RA pressure | -0.33 | 0.01 | <0.001 |  |  |  |
| Mean PA pressure | -0.68 | 0.01 | <0.001 |  |  |  |
| Mean PCWP pressure | -0.44 | 0.01 | <0.001 |  |  |  |
| Cardiac Output | 0.52 | 0.01 | <0.001 |  |  |  |
| PVR | -0.62 | 0.01 | <0.001 |  |  |  |

Association of clinical variables with PAC was evaluated in age- and sex-adjusted linear regression models. Beta coefficient represents change in PAC per 1SD change in clinical variable (or presence vs absence for dichotomous variables). Partial correlation of RHC measures and PAC was examined with Pearson partial correlation coefficients, adjusted for age and sex. \*NT-proBNP available for 3,598 (45%) patients; excluded from stepwise model.

Hemodynamic correlates included mean RA, mean PA, mean PCWP, and PVR, which were all inversely associated with PAC, and cardiac output, which was positively associated with PAC (partial correlation coefficient P<0.001 for all).

### Association of lower PAC with adverse clinical outcomes

We observed 2,894 deaths and 1,925 HF hospitalizations over a median follow up time of 7.4 years. The Kaplan-Meier curves for overall mortality and HF hospitalization demonstrate progressively worse outcomes with lower PAC quartile (log rank p < 0.001 for both outcomes) **Figure 2**).

**Figure 2:**
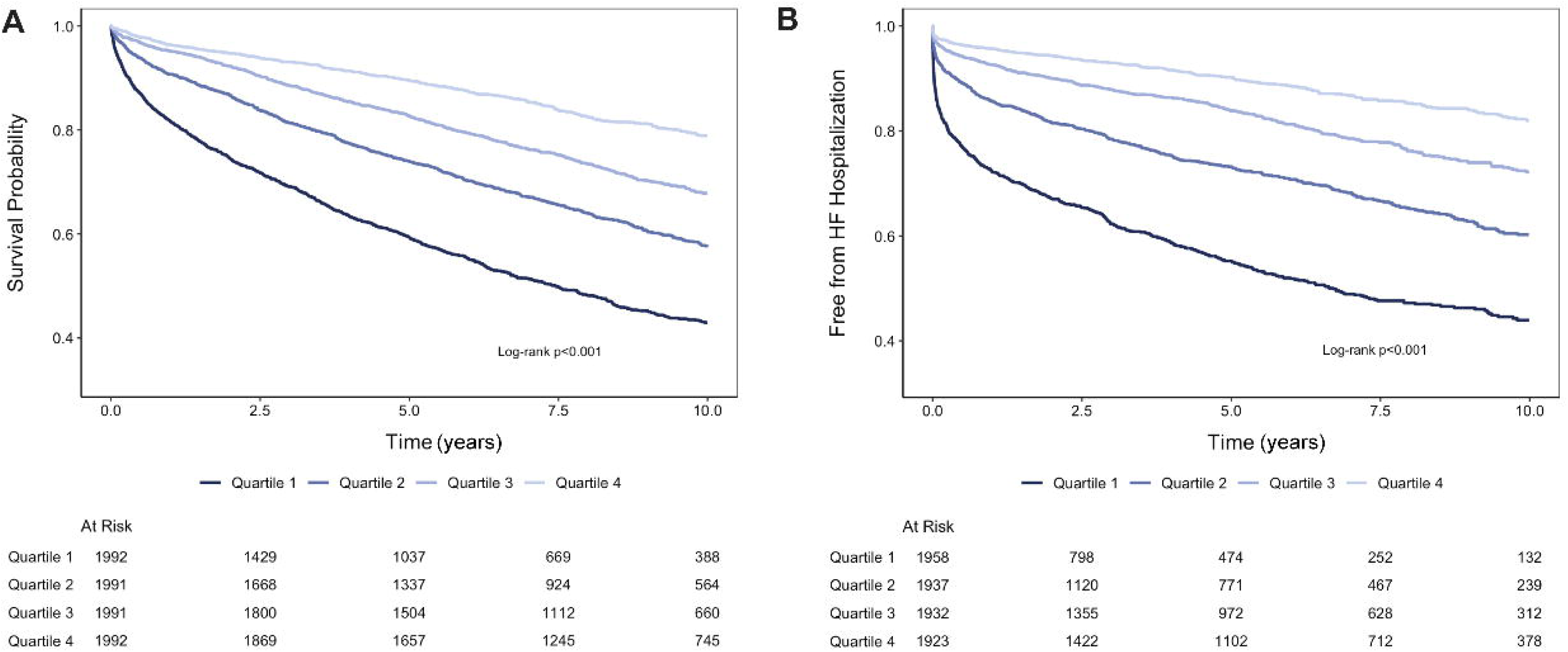
Kaplan Meier curves demonstrating (A) survival and (B) freedom from heart failure hospitalization by PAC quartile. Number at risk is listed below the plot. HF: heart failure.

In age and sex-adjusted Cox models, a 1-SD higher PAC (equivalent to 5.65 mL/mmHg change in PAC) was associated with a 45% lower risk of all-cause death (HR 0.55, 95% CI 0.52, 0.58, p<0.001) and a 49% lower risk of HF hospitalization (HR 0.51, 95% CI 0.48, 0.54, p<0.001) (**Supplementary Table 1**). These associations persisted in both the multivariable clinical model for mortality (HR 0.59, 95% CI 0.56, 0.63, p<0.001) and HF hospitalization risk (HR 0.56, 95% CI 0.52, 0.59, p<0.001). Addition of PVR to the multivariable model also demonstrated a significant association: a 1-SD higher PAC was associated with a 37% lower risk of all-cause death (HR 0.63, 95% CI 0.59, 0.68, p <0.001) and 42% lower risk of HF hospitalization (HR 0.58, 95% CI 0.54, 0.62, p <0.001).

### Performance of PAC vs other measures of pulmonary vascular dysfunction

We examined performance metrics of a clinical risk prediction model (including age, sex, BMI, history of hypertension, diabetes mellitus, previous myocardial infarction, chronic HF, chronic kidney disease, OSA/OHS, and chronic lung disease) of all-cause mortality with and without the addition of PAC and other measures of pulmonary vascular function (**Table 4**). The base clinical model yielded a c-statistic of 0.680 (95% CI 0.669, 0.690). There was a significant improvement in model discrimination with the addition of PAC (delta c-statistic 0.027, 95% CI 0.021, 0.033, p<0.001). Additionally, PAC provided incremental information when added to the clinical model with PVR (delta c-statistic 0.013, 95% CI 0.009, 0.017, p<0.001). In risk prediction models of HF hospitalization, the base clinical model demonstrated a c-statistic of 0.676 (95% CI 0.664, 0.688). Addition of PAC significantly improved model discrimination (delta c-statistic 0.021, 95% CI 0.008, 0.034, p=0.001), with similar incremental value as the addition of PVR (delta c-statistic 0.021, 95% CI 0.010, 0.032, p<0.001).

### Association of PAC with adverse clinical outcomes in PH

Across the entire cohort, 63% met hemodynamic criteria for PH. PAC was lower among those with PH (2.6 mL/mmHg, IQR 1.8, 3.7) compared to without PH (4.5 mL/mmHg, IQR 3.5, 5.9, p<0.001). Among those with PH, we observed 2,217 deaths and 1,571 HF hospitalizations over a median follow up time of 6.4 years, and among those without PH we observed 677 deaths and 254 HF hospitalizations over a median follow up time of 8.9 years. Kaplan-Meier curves for overall mortality in those with and without PH demonstrate progressively worse outcomes with lower PAC quartile for both groups (log rank p < 0.001 for each) (**Figure 3**).

**Figure 3:**
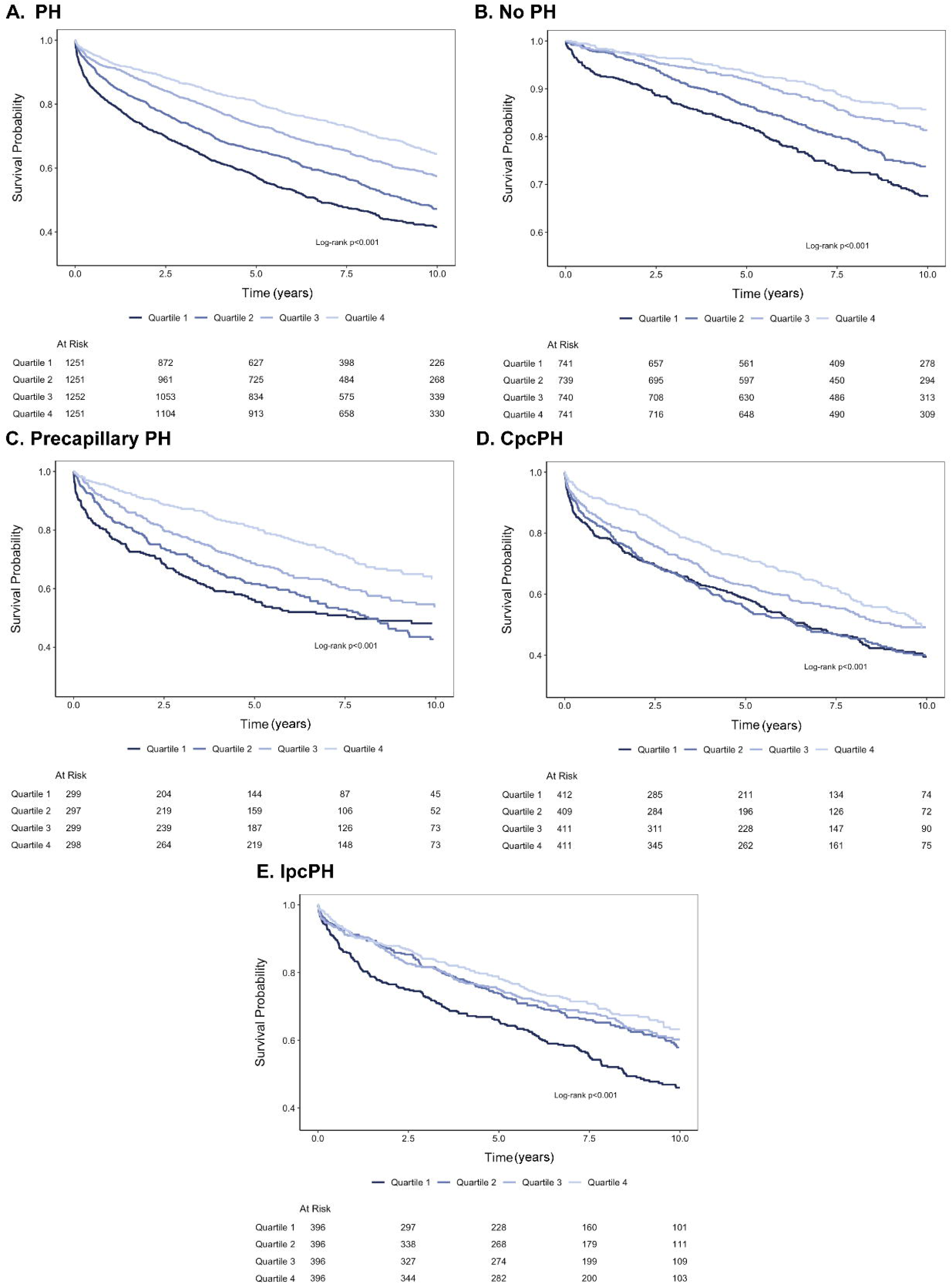
Association of PAC quartile with all-cause mortality by presence and absence of pulmonary hypertension (PH) (A, PH; B, no PH) and by PH hemodynamic subgroup: C, precapillary; D, isolated postcapillary (IpcPH); E, combined pre- and post-capillary (CpcPH). Number at risk is listed below the plot.

In multivariable Cox models, among those with PH, a 1 SD higher PAC was associated with a 31% lower mortality risk (HR 0.69, 95% CI 0.64, 0.74, p < 0.001) and among those without PH, a 1 SD higher PAC was also associated with a 26% lower mortality risk (HR 0.74, 95% CI 0.66, 0.84, p < 0.001) (**Table 3**, **Figure 4**). After addition of PVR to the clinical model, similar results were observed (PH: HR 0.72, 95% CI 0.66, 0.79, p<0.001; no PH: HR 0.72, 95% CI 0.64, 0.82, p<0.001). There was similarly a significant inverse association between PAC and HF hospitalization risk in those with (HR 0.65, 95% CI 0.60, 0.70) and without PH (HR 0.73, 95% CI 0.61, 0.87) in the multivariable model.

**Figure 4:**
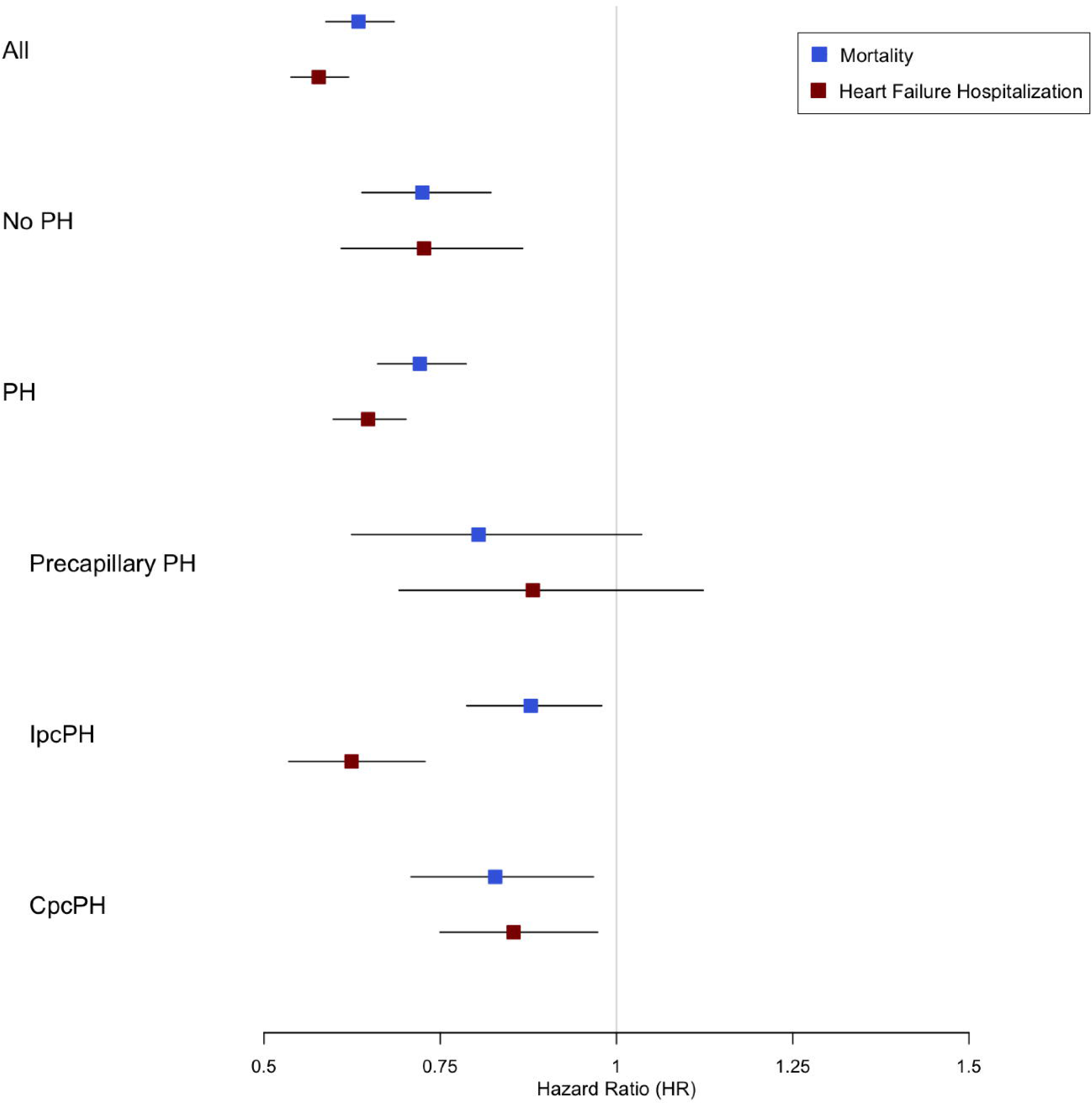
Association of PAC with mortality (top) and heart failure (HF) hospitalization (bottom) by pulmonary hypertension (PH) subgroup in multivariable-adjusted analyses. Models adjusted for age, sex, body mass index, history of hypertension, diabetes mellitus, previous myocardial infarction, chronic HF, chronic kidney disease, OSA/OHS, chronic lung disease, and pulmonary vascular resistance. IpcPH: isolated post-papillary pulmonary hypertension; CpcPH: combined pre- and postcapillary pulmonary hypertension.

**Table 3:**
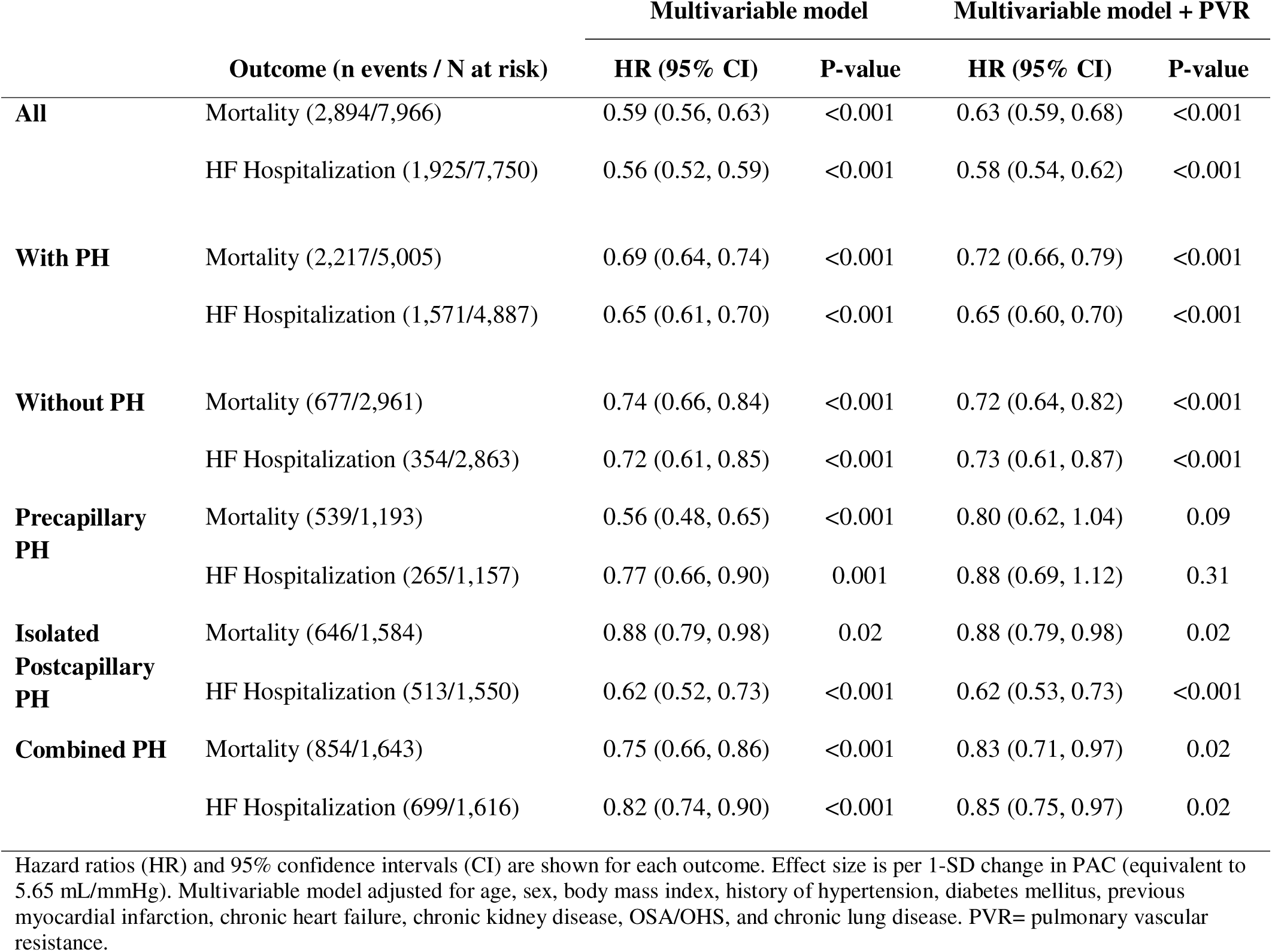
Association of PAC with clinical outcomes across the entire sample, and stratified by pulmonary hypertension (PH) status.

|  | Outcome (n events / N at risk) | Multivariable model |  | Multivariable model + PVR |  |
| --- | --- | --- | --- | --- | --- |
|  |  | HR (95% CI) | P-value | HR (95% CI) | P-value |
| **All** | Mortality (2,894/7,966) | 0.59 (0.56, 0.63) | <0.001 | 0.63 (0.59, 0.68) | <0.001 |
|  | HF Hospitalization (1,925/7,750) | 0.56 (0.52, 0.59) | <0.001 | 0.58 (0.54, 0.62) | <0.001 |
| **With PH** | Mortality (2,217/5,005) | 0.69 (0.64, 0.74) | <0.001 | 0.72 (0.66, 0.79) | <0.001 |
|  | HF Hospitalization (1,571/4,887) | 0.65 (0.61, 0.70) | <0.001 | 0.65 (0.60, 0.70) | <0.001 |
| **Without PH** | Mortality (677/2,961) | 0.74 (0.66, 0.84) | <0.001 | 0.72 (0.64, 0.82) | <0.001 |
|  | HF Hospitalization (354/2,863) | 0.72 (0.61, 0.85) | <0.001 | 0.73 (0.61, 0.87) | <0.001 |
| **Precapillary PH** | Mortality (539/1,193) | 0.56 (0.48, 0.65) | <0.001 | 0.80 (0.62, 1.04) | 0.09 |
|  | HF Hospitalization (265/1,157) | 0.77 (0.66, 0.90) | 0.001 | 0.88 (0.69, 1.12) | 0.31 |
| **Isolated Postcapillary PH** | Mortality (646/1,584) | 0.88 (0.79, 0.98) | 0.02 | 0.88 (0.79, 0.98) | 0.02 |
|  | HF Hospitalization (513/1,550) | 0.62 (0.52, 0.73) | <0.001 | 0.62 (0.53, 0.73) | <0.001 |
| **Combined PH** | Mortality (854/1,643) | 0.75 (0.66, 0.86) | <0.001 | 0.83 (0.71, 0.97) | 0.02 |
|  | HF Hospitalization (699/1,616) | 0.82 (0.74, 0.90) | <0.001 | 0.85 (0.75, 0.97) | 0.02 |

Hazard ratios (HR) and 95% confidence intervals (CI) are shown for each outcome. Effect size is per 1-SD change in PAC (equivalent to 5.65 mL/mmHg). Multivariable model adjusted for age, sex, body mass index, history of hypertension, diabetes mellitus, previous myocardial infarction, chronic heart failure, chronic kidney disease, OSA/OHS, and chronic lung disease. PVR= pulmonary vascular resistance.

**Table 4:** Performance of PAC versus other measures of pulmonary vascular dysfunction.

| Outcome | C statistic | 95% CI | Delta C statistic* | 95% CI | p-value |
| --- | --- | --- | --- | --- | --- |
| **Mortality** |  |  |  |  |  |
| Clinical | 0.680 | 0.669, 0.690 |  |  |  |
| Clinical model + PAC | 0.704 | 0.694, 0.714 | 0.027 | 0.021, 0.033 | <0.001 |
| Clinical model + PVR | 0.694 | 0.683, 0.704 | 0.014 | 0.008, 0.020 | <0.001 |
| Clinical model + PAC + PVR | 0.707 | 0.697, 0.716 | 0.027 | 0.021, 0.033 | <0.001 |
|  |  |  | 0.013<sup>†</sup> | 0.009, 0.017 | <0.001 |
| Clinical model + RC | 0.680 | 0.667, 0.687 | 0 | -0.002, 0.002 | 0.58 |
| **HF hospitalization** |  |  |  |  |  |
| Clinical | 0.676 | 0.664, 0.688 |  |  |  |
| Clinical model + PAC | 0.711 | 0.698, 0.730 | 0.021 | 0.008, 0.034 | 0.001 |
| Clinical model + PVR | 0.697 | 0.682, 0.712 | 0.021 | 0.010, 0.032 | 0.001 |
| Clinical model + PAC + PVR | 0.698 | 0.677, 0.718 | 0.022 | 0.008, 0.036 | 0.001 |
|  |  |  | 0.001 | -0.009, 0.007<sup>†</sup> | 0.78 |
| Clinical model + RC | 0.675 | 0.658, 0.692 | -0.001 | -0.003, -0.001 | 0.191 |

Clinical model adjusted for age, sex, body mass index, history of hypertension, diabetes mellitus, previous myocardial infarction, chronic heart failure, chronic kidney disease, obesity hypoventilation syndrome/obstructive sleep apnea, and chronic lung disease.
\*Delta-c statistic comparing to base clinical model, unless otherwise noted; <sup>†</sup>Delta C statistic comparing to clinical model + PVR; PAC: pulmonary artery compliance; PVR: pulmonary vascular resistance; RC: resistance-compliance time constant.

PAC significantly differed between PH hemodynamic subgroups groups (p<0.001): pre-capillary PH: 2.5 mL/mmHg (IQR 1.7, 3.2), IpcPH: 3.2 mL/mmHg (IQR 2.4, 4.3), and CpcPH PH: 1.78 mL/mmHg (IQR 1.3, 2.4). Demographics and hemodynamics by PH subtype are reported in Supplementary Table 2.

Among those with precapillary PH, we observed 539 deaths over a median follow up time of 6.1 years; IpcPH 646 deaths over a median follow up time of 7.0 years; and CpcPH 854 deaths over a median follow up time of 5.5 years. Kaplan Meier curves for mortality by PAC quartile for each hemodynamic PH subgroup are shown in **Figure 3**. Log-rank pairwise comparisons demonstrated that among those with pre-capillary PH and CpcPH, mortality was similar in Quartile 1 and 2, yet significantly differed between all other quartile pairings. Among those with IpcPH, mortality was significantly higher in Quartile 1 compared to the remaining quartiles, which had similar mortality.

In multivariable-adjusted Cox models of individuals with pre-capillary PH, a 1-SD higher PAC was associated with a 44% lower hazard of mortality (HR 0.56, 95% CI 0.48, 0.65, p<0.001) and a 23% lower risk of HF hospitalization (HR 0.77, 95% CI 0.66, 0.90, p=0.001) (**Table 3**, **Figure 4**). Similarly, PAC was associated with mortality and HF hospitalization among those with CpcPH (mortality: HR 0.75, 95% CI 0.66, 0.86; HF hospitalization: HR 0.82, 95% CI 0.74, 0.90) and IpcPH (mortality: HR 0.88, 95% CI 0.79, 0.98; HF hospitalization: HR 0.62, 95% CI 0.52, 0.73). With addition of PVR to the multivariable model, PAC remained significantly associated with mortality and HF hospitalization among those with CpcPH (mortality: HR 0.83, 95% CI 0.71, 0.97, p=0.02; HF hospitalization: HR 0.85, 95% CI 0.75, 0.97, p=0.02) and IpcPH (mortality: HR 0.88, 95% CI 0.79, 0.98, p=0.02; HF hospitalization: HR 0.62, 95% CI 0.53, 0.73, p<0.001), yet among those with precapillary PH, PAC was no longer significantly associated with mortality and HF hospitalization (mortality: HR 0.80, 95% CI 0.62, 1.04, p=0.09; HF hospitalization: HR 0.88, 95% CI 0.69, 1.12, p=0.31).

### RC Time differs between PH subgroups

The RC time constant, the product of PAC and PVR, significantly differed by PH subgroup, shortest in the IpcPH subgroup (0.23 seconds, IQR 0.16, 0.32), compared to the pre-capillary (0.52 seconds, IQR 0.42, 0.63) and CpcPH subgroups (0.36 seconds, IQR 0.28, 0.45; p<0.001 for all between-group differences) (**Figure 5A, Supplementary Table 2**). There was an inverse relationship between PAC and PVR across all hemodynamic subtypes (**Figure 5B**). For the same PVR, PAC was lower among those with CpcPH versus those with precapillary PH, and those with IpcPH had a lower PAC than those without PH.

**Figure 5:**
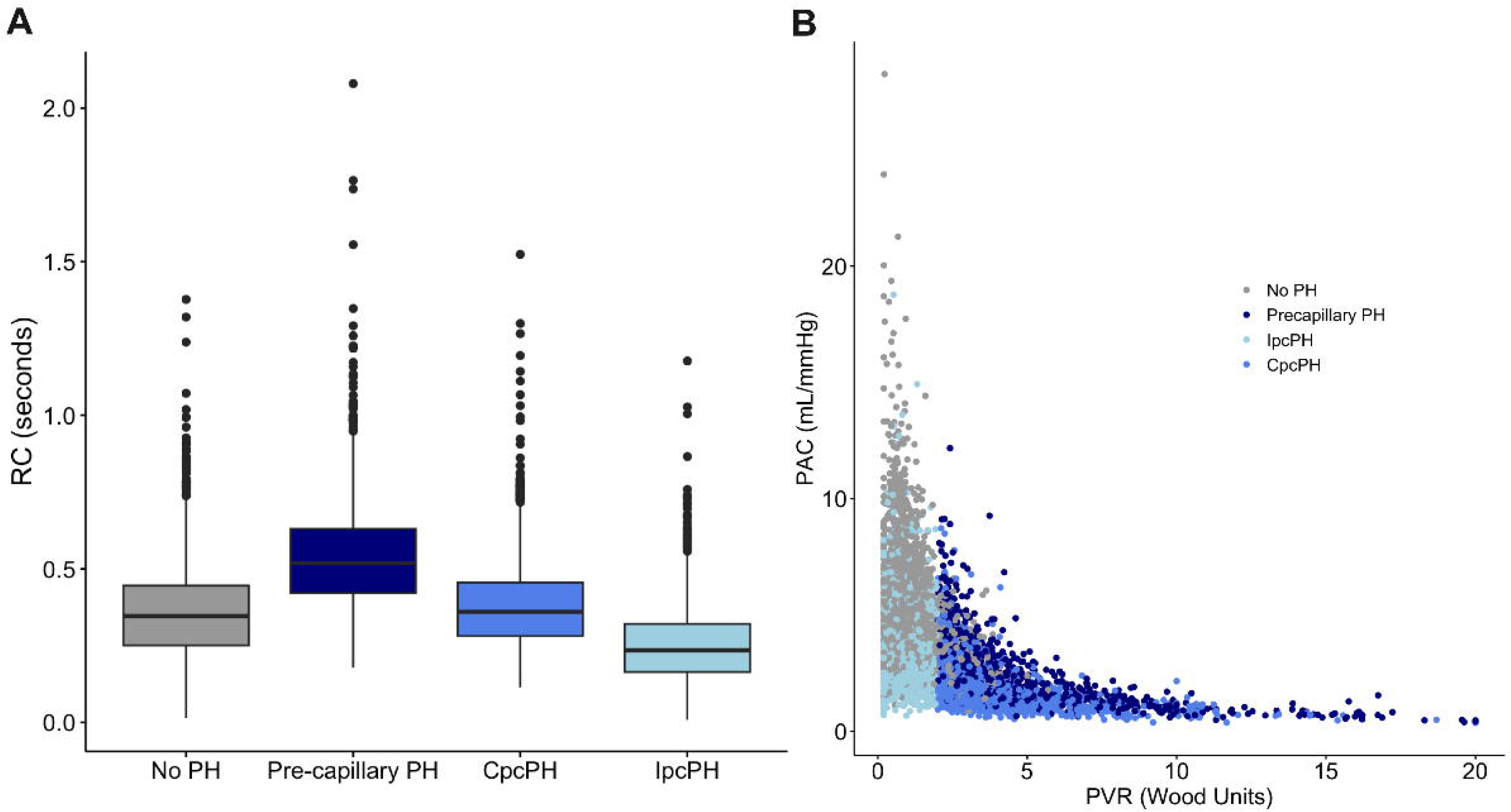
Distribution of RC time differs by pulmonary hypertension (PH) subgroup (panel A, all pairwise comparisons p<0.001), with two components of RC time demonstrating logarithmic relationship between PVR and PAC, with distinct clusters by PH subgroup indicated by colors. In panel A, all between group relationships demonstrate statistical significance with p<0.001. IpcPH: isolated post-papillary pulmonary hypertension; CpcPH: combined pre- and postcapillary pulmonary hypertension; PAC pulmonary artery compliance; PVR: pulmonary vascular resistance.

## Discussion

In this broad single-site sample of individuals undergoing invasive hemodynamic evaluation with RHC, we found that worse PAC, a measure of pulsatile RV afterload, was associated with greater all-cause mortality and HF hospitalizations irrespective of the presence or absence of PH and across all PH hemodynamic subtypes. Further, among those with IpcPH and CpcPH, PAC remained independently associated with worse prognosis even after accounting for PVR, a measure of resistive RV afterload. Lastly, addition of PAC to a clinical risk model improved discrimination with respect to mortality even beyond PVR. Taken together, our data extend the findings of prior studies focused on specific PH samples, and demonstrate that PAC is associated with adverse clinical outcomes across a broad sample including individuals irrespective of PH status. These findings underscore the importance of examining the future clinical applications of PAC beyond risk assessment, including potential use in diagnosis and monitoring response to therapeutics.

Contemporary PH hemodynamic classification, namely distinguishing between IpcPH and CpcPH, and risk stratification tools, such as the Registry to Evaluate Early and Long-Term PAH Disease Management (REVEAL) 2.0, largely utilize PVR as the hemodynamic metric to assess disease severity.^2,13^ Given this only reflects the static component of RV afterload, PAC has emerged as an important area of study. Determined by the ratio of stroke volume to pulmonary artery pulse pressure, PAC reflects distensibility of the pulmonary circulation and the pulsatile component of right ventricular afterload.^14^ Lower PAC has been demonstrated to be associated with adverse outcomes among multiple subgroups, including in a large network analysis of those with mild precapillary PH and in smaller cohorts of patients with pulmonary arterial hypertension, chronic thromboembolic pulmonary hypertension, and PH associated with left heart disease.^8–10^ We extend these findings to a heterogeneous sample of individuals presenting for RHC and highlight the prognostic significance of PAC in individuals with and without PH. In this large sample, PAC was associated with greater risk of death and HF hospitalization, independent of potential clinical confounders. Notably, among those with and without PH, the association of PAC with adverse events remained significant independent of PVR. Further, when added to clinical variables, the discrimination of risk model for mortality incorporating both PAC and PVR was superior to the clinical model with PVR alone. This underscores the added prognostic value with consideration of both the static and pulsatile components of RV afterload in assessing disease severity.

PAC was significantly associated with mortality and HF hospitalization across all hemodynamic PH subtypes. After addition of PVR to the multivariable model, PAC remained significantly associated with both outcomes in those with IpcPH and CpcPH, yet not in those with precapillary PH. This may be explained by the larger influence of PAC on afterload in post-capillary disease due to elevated left-sided pressures.^6,15^ Prior studies have demonstrated that the inverse hyperbolic relationship between PAC and PVR, the RC time constant, is shifted down and to the left in cases of elevated PCWP.^6,16^ Accordingly, for every resistance, there is a lower compliance among those with post-capillary disease compared to pre-capillary disease.^6^ Thus in IpcPH and CpcPH, both states of elevated left sided pressure, PAC may remain prognostically important in addition to PVR due to a greater influence on RV afterload. This is further illustrated by the fact precapillary and CpcPH groups had similar PVR, yet PAC provided additional prognostic value only in the latter cohort. Our data demonstrate that PAC provides a supplemental measure of the dynamic component of RV afterload that is particularly prognostically important among those with elevated left-sided pressures.

Among our cohort, invasive assessment of PAC was associated with mortality and HF hospitalization even in patients without PH. While this has not been explicitly studied to our knowledge, this finding aligns with a prior study of an elderly, community-based cohort without heart failure, in which low PAC measured by echocardiogram was found to be associated with a composite of heart failure and death after adjusting for demographic and echocardiographic parameters.^17^ Given the logarithmic relationship between PAC and PVR, as demonstrated in Figure 4, PAC is variable at lower PVR, yet at higher PVR, reflecting overt disease states, there is minimal change in PAC with change in PVR.^3^ This suggests that PAC may serve as a marker for “pre-disease” states, identifying early or subclinical stages of pulmonary vascular remodeling and/or heart failure with preserved ejection fraction.

In examination of clinical and hemodynamic correlates of PAC, we found that lower PAC was associated with older age and female sex. This is in accordance with prior studies which demonstrated lower PAC in women compared to men at diagnosis.^18,19^ The “sex paradox” in PAH, in which women are at higher risk for PAH yet have higher survival than men, is postulated to be related to the interaction between hormones, RV function, and comorbidity burden.^19^ The lower PAC at diagnosis may be a reflection of this, although future gender-based studies examining PAC across the disease course are warranted. We also found that lower PAC was associated with cardiopulmonary comorbidities, including chronic HF and chronic lung disease, and other hemodynamic indicators of disease severity, including higher PVR and lower cardiac output. Interestingly, a higher PAC was associated with a higher BMI, aligning with prior studies, likely driven by the higher cardiac output in individuals with obesity.^20^ The association of higher PAC with systemic hypertension has not been well-described. While they share certain pathological mechanisms, this underscores that changes in compliance in the pulmonary circulation cannot be extrapolated to similar changes in the systemic circulation due to differential distribution of compliance between the two systems.^21,22,23^ While our findings highlight the complex interplay between PAC and clinical comorbidities, they support an association of PAC with adverse outcomes even after accounting for these potential clinical confounders.

There are several limitations in our analysis. Our study included individuals presenting for both outpatient hospital-based clinically-indicated RHC which may introduce selection or referral bias, limiting generalizability to other samples. Further, unmeasured confounding and the observational nature of the study precludes causal inferences. HF hospitalization was determined via ICD and CPT codes which may have misclassified cases, although we would expect this to uniformly affect our cohort regardless of PAC. We did not have available data for World Health Organization PH group determination, limiting inferences on exact PH etiology, or complete information required for calculation of existing prognostic risk scores, including REVEAL 2.0.^13^ Finally, examination of single RHC instances did not allow for analysis of treatment effects of PAC.

In conclusion, we demonstrate the prognostic significance of PAC in a broad hospital-based sample of individuals across the spectrum of cardiopulmonary disease. We found that lower PAC was associated with greater risk of all-cause death and HF hospitalization, irrespective of presence or absence of PH or PH subtype. Further, PAC as a measure of pulsatile RV afterload remained independently associated with worse prognosis even after accounting for PVR as a measure of resistive RV afterload in patients with post-capillary disease. Our findings highlight the additive prognostic value of PAC that may indicate early pulmonary vascular disease even when PVR is not yet abnormal. Further studies are needed to examine the potential of PAC to identify high-risk individuals for tailored preventive or therapeutic approaches in pulmonary vascular disease.

## Data Availability

All data produced in the present study are available upon reasonable request to the authors

## Acknowledgements

L.A.M and J.E.H designed the study. J.E.H, J.N.M, M.H.P, and E.V.P assisted with database formation. L.A.M and J.P performed the analysis. L.A.M and S.M.N compiled the manuscript. L.B.K, J.N.M, M.H.P, E.B.P, D.F., N.S, and C.K provided critical feedback and framing.

## Abbreviations

PH: pulmonary hypertension
PAC: pulmonary artery compliance
PVR: pulmonary vascular resistance
RV: right ventricular
RC: resistance compliance time constant
RHC: right heart catheterization
MI: myocardial infarction
HF: heart failure
BMI: body mass index
OSA/OHS: obstructive sleep apnea/ obesity hypoventilation syndrome
RA: right atrial
PA: pulmonary artery
PCWP: pulmonary capillary wedge pressure
IpcPH: isolated post-capillary pulmonary hypertension
CpcPH: combined pre- and post-capillary pulmonary hypertension
WU: Wood units
NT-proBNP: N-Terminal Pro-B-Type Natriuretic Peptide
REVEAL: Registry to Evaluate Early and Long-Term PAH Disease Management

