## Supplemental Tables for "Association of Pulmonary Artery Compliance and Adverse Cardiac Events"

**Supplementary Table 1:** Age- and sex- adjusted association of PAC with clinical outcomes across the entire sample, and stratified by pulmonary hypertension (PH) status

|  |  | **Age/sex adjusted model** | |
| --- | --- | --- | --- |
|  | **Outcome (n events / N at risk)** | **HR (95% CI)** | **P-value** |
| **All** | Mortality (2,894/7,966) | 0.55 (0.52, 0.58) | <0.001 |
|  | HF Hospitalization (1,925/7,750) | 0.51 (0.48, 0.54) | <0.001 |
| **With PH** | Mortality (2,217/5,005) | 0.64 (0.59, 0.68) | <0.001 |
|  | HF Hospitalization (1,571/4,887) | 0.61 (0.58, 0.66) | <0.001 |
| **Without PH** | Mortality (677/2,961) | 0.67 (0.60, 0.76) | <0.001 |
|  | HF Hospitalization (354/2,863) | 0.64 (0.54, 0.75) | <0.001 |
| **Precapillary PH** | Mortality (539/1,193) | 0.52 (0.45, 0.60) | <0.001 |
|  | HF Hospitalization (265/1,157) | 0.81 (0.70, 0.93) | 0.004 |
| **Isolated Postcapillary PH** | Mortality (646/1,584) | 0.84 (0.75, 0.93) | 0.001 |
|  | HF Hospitalization (513/1,550) | 0.59 (0.51, 0.68) | <0.001 |
| **Combined PH** | Mortality (854/1,643) | 0.71 (0.62, 0.81) | <0.001 |
|  | HF Hospitalization (699/1,616) | 0.78 (0.71, 0.86) | <0.001 |

Hazard ratios (HR) and 95% confidence intervals (CI) are shown for each outcome. Effect size is per 1-SD change in PAC. PH: pulmonary hypertension.

**Supplementary Table 2:** Demographics, comorbidities, echocardiographic data, and hemodynamics by pulmonary hypertension subgroup

| **Characteristic** | | **No PH**  N = 2,961 | | **Pre-capillary PH**  N = 1,193 | | **IpcPH**  N = 1,584 | | **CpcPH**  N = 1,643 |
| --- | --- | --- | --- | --- | --- | --- | --- | --- |
| Age | | 61 (14) | | 64 (12) | | 63 (12) | | 65 (11) |
| Sex | |  | |  | |  | |  |
| Female | | 1,186 (40%) | | 579 (49%) | | 467 (29%) | | 699 (43%) |
| Male | | 1,775 (60%) | | 614 (51%) | | 1,117 (71%) | | 944 (57%) |
| BMI | | 28 (6) | | 28 (7) | | 32 (8) | | 30 (7) |
| Hypertension | | 1,669 (56%) | | 610 (51%) | | 1,046 (66%) | | 1,001 (61%) |
| Chronic heart failure | | 573 (19%) | | 319 (27%) | | 669 (42%) | | 852 (52%) |
| Previous MI | | 455 (15%) | | 190 (16%) | | 335 (21%) | | 388 (24%) |
| Diabetes | | 503 (17%) | | 243 (20%) | | 478 (30%) | | 489 (30%) |
| Recent smoker | | 132 (8.0%) | | 65 (9.9%) | | 91 (9.8%) | | 121 (12%) |
| Chronic kidney disease | | 36 (1.2%) | | 22 (1.8%) | | 71 (4.5%) | | 81 (4.9%) |
| Chronic lung disease | | 389 (13%) | | 268 (22%) | | 242 (15%) | | 294 (18%) |
| OSA/ OHS | | 279 (9.1%) | | 171 (14%) | | 316 (20%) | | 248 (15%) |
| NT-proBNP | | 608 (157, 1,919) | | 1,686 (585, 4,155) | | 1,860 (676, 5,007) | | 3,029 (1,424, 6,978) |
| Ejection fraction (%) | | 64 (49, 70) | | 63 (43, 71) | | 55 (31, 67) | | 44 (25, 64) |
| RVSP | | 36 (31, 42) | | 51 (41, 65) | | 45 (37, 53) | | 53 (44, 63) |
| Mean RA pressure | | 4.0 (2.0, 6.0) | | 6.0 (4.0, 9.0) | | 10.0 (7.0, 13.0) | | 11.0 (8.0, 15.0) |
| Systolic PA pressure | | 26 (23, 30) | | 41 (36, 52) | | 42 (37, 50) | | 55 (47, 66) |
| Diastolic PA pressure | | 9 (6, 11) | | 16 (13, 21) | | 19 (15, 23) | | 24 (19, 29) |
| Mean PA pressure | | 16 (14, 18) | | 26 (23, 32) | | 28 (25, 33) | | 37 (32, 44) |
| Mean PCWP Pressure | | 9 (7, 11) | | 11 (9, 13) | | 21 (18, 26) | | 22 (18, 27) |
| Cardiac Output (L/min) | | 5.21 (4.35, 6.24) | | 4.65 (3.90, 5.58) | | 5.40 (4.49, 6.65) | | 4.19 (3.44, 5.15) |
| PVR (Wood Units) | | 1.25 (0.85, 1.71) | | 3.25 (2.50, 4.68) | | 1.29 (0.88, 1.61) | | 3.11 (2.45, 4.26) |
| PAC (mL/mmHg) | | 4.49 (3.51, 5.94) | | 2.48 (1.75, 3.23) | | 3.18 (2.37, 4.34) | | 1.78 (1.30, 2.39) |
| PAC Quartile |  | |  | |  | |  | |
| Quartile 1 | | 0 (0%) | | 7 (0.6%) | | 0 (0%) | | 8 (0.5%) |
| Quartile 2 | | 1 (<0.1%) | | 80 (6.7%) | | 10 (0.6%) | | 204 (12%) |
| Quartile 3 | | 47 (1.6%) | | 298 (25%) | | 188 (12%) | | 757 (46%) |
| Quartile 4 | | 2,913 (98%) | | 808 (68%) | | 1,386 (88%) | | 674 (41%) |
| RC time | | 0.35 (0.25, 0.45) | | 0.52 (0.42, 0.63) | | 0.23 (0.16, 0.32) | | 0.36 (0.28, 0.45) |
| Heart rate | | 67 (59, 77) | | 73 (63, 85) | | 71 (62, 82) | | 74 (65, 86) |
| MI: myocardial infarction; OSA: obstructive sleep apnea; OHS: obesity hypoventilation syndrome; RA: right atrial; PA: pulmonary artery; PCWP: pulmonary capillary wedge pressure; PVR: pulmonary vascular resistance; PAC: pulmonary artery compliance; RC: RC time constant.  Median and interquartile range for continuous variable and count and percent for categorical variables, unless otherwise noted  ^*^ Mean (standard deviation) | | | | | | | | |
